# Sensitivity of the Elecsys Anti-SARS-CoV-2 immunoassay as an aid in determining previous exposure to SARS-CoV-2

**DOI:** 10.1101/2021.02.11.21250290

**Authors:** Johannes Kolja Hegel, Elena Riester, Christopher M. Rank, Florina Langen, Tina Laengin, Peter Findeisen

## Abstract

**Background:** The Elecsys® Anti-SARS-CoV-2 electrochemiluminescence immunoassay (Roche Diagnostics International Ltd) was developed for the *in vitro* qualitative detection of antibodies to SARS-CoV-2. We evaluated the sensitivity of the Elecsys Anti-SARS-CoV-2 immunoassay in samples from a diverse cross-section of patients across multiple sites and compared results against commercially available comparators.

**Methods:** Sensitivity of the Elecsys Anti-SARS-CoV-2 immunoassay was evaluated using anonymised, frozen, residual single and sequential serum and plasma samples from patients with polymerase chain reaction (PCR)-confirmed SARS-CoV-2 infection. Point estimates and 95% confidence intervals (CIs) were calculated and method comparisons performed versus the following comparator assays: Euroimmun Anti-SARS-CoV-2 IgG, Abbott ARCHITECT SARS-CoV-2 IgG, Siemens ADVIA Centaur SARS-CoV-2 Total, and YHLO iFlash SARS-CoV-2 IgG and IgM.

**Results:** Overall sensitivity for the Elecsys Anti-SARS-CoV-2 immunoassay in 219 samples drawn ≥14 days post-PCR confirmation was 93.6% (95% CI 89.5–96.5). Across the three study sites, sensitivity in samples drawn ≥14 days post-PCR confirmation ranged from 85.7–98.9%. Sensitivity was significantly higher for the Elecsys Anti-SARS-CoV-2 immunoassay compared with the YHLO iFlash SARS-CoV-2 IgM assay for samples drawn ≥14 days post-PCR confirmation (86.3% [95% CI 76.7–92.9] versus 33.8% [95% CI 23.6–45.2]). Both Siemens ADVIA Centaur SARS-CoV-2 Total and YHLO iFlash SARS-CoV-2 IgG assays had a significantly higher sensitivity compared with the Elecsys Anti-SARS-CoV-2 immunoassay for samples drawn ≥14 days post-PCR confirmation (95.1% [95% CI 87.8–98.6] versus 85.2% [95% CI 75.6–92.1]; 93.8% [95% CI 86.0–97.9] versus 86.3% [95% CI 76.7–92.9]). Differences in sensitivity between the Elecsys Anti-SARS-CoV-2 immunoassay and the Euroimmun Anti-SARS-CoV-2 IgG (90.3% [95% CI 83.7–94.9] versus 95.2% [95% CI 89.8–98.2]) and Abbott ARCHITECT SARS-CoV-2 IgG (84.8% [95% CI 75.0–91.9] versus 87.3% [95% CI 78.0–93.8]) assays for samples drawn ≥14 days post-PCR confirmation were not significant.

**Conclusions:** The Elecsys Anti-SARS-CoV-2 immunoassay demonstrated high sensitivity in samples collected ≥14 days post-PCR confirmation of SARS-CoV-2 infection, and comparable sensitivity to several commercially available comparator assays across multiple sites, supporting the use of this assay as a tool to aid in determination of previous exposure to SARS-CoV-2.

**Required information for submission system:** *Ethical guidelines:* The study was conducted in accordance with the study protocol provided by Roche Diagnostics and in accordance with the principles of the Declaration of Helsinki. All human samples utilised at the three study sites in Germany (Augsburg, Heidelberg, Berlin) were anonymised, frozen, residual samples, therefore no ethical approval or waiver was required in accordance with local legislation from ZEKO (Central Ethics Commission at the German Medical Association). A statement was obtained from the Ethics Committee of the Landesärztekammer Bayern confirming that there are no objections to the coherent use of anonymised residual samples.

*Research reporting guidelines:* Please see separate STARD checklist.

*Data availability statement:* Qualified researchers may request access to individual patient level data through the clinical study data request platform (https://vivli.org/). Further details on Roche’s criteria for eligible studies are available here: https://vivli.org/members/ourmembers/. For further details on Roche’s Global Policy on the Sharing of Clinical Information and how to request access to related clinical study documents, see here: https://www.roche.com/research_and_development/who_we_are_how_we_work/clinical_trials/our_commitment_to_data_sharing.htm

## Introduction

As of 17^th^ December 2020, there have been more than 72.5 million confirmed cases worldwide of the novel severe acute respiratory syndrome coronavirus 2 (SARS-CoV-2), including over 1.6 million deaths.^1^ SARS-CoV-2 is the virus responsible for the coronavirus disease 2019 (COVID-19) pandemic, originating from Wuhan, China.^2,3^ SARS-CoV-2 is an enveloped, non-segmented, single-stranded RNA virus that shares similarities with other coronaviruses in the expression of its genome, which encodes 16 non-structural proteins and four structural proteins, known as the spike (S), envelope (E), membrane (M) and nucleocapsid (N) antigens.^4,5^ Individuals infected with SARS-CoV-2 may exhibit a range of respiratory symptoms, including a persistent cough and shortness of breath, in addition to fever and fatigue.^2,6^ Although up to 80% of infections are mild or asymptomatic, 15% are severe, requiring oxygen, and 5% are critical, requiring ventilation.^6^ Symptomatic and pre-symptomatic transmission of SARS-CoV-2, occurring via contact with infected respiratory droplets and contaminated surfaces, is thought to play a greater role in the spread of the virus than asymptomatic transmission.^7,8^

Reverse transcriptase polymerase chain reaction (RT-PCR) is the current gold standard for detection of SARS-CoV-2 RNA in sputum gathered from patient nasopharyngeal swabs, which typically have high viral titres during the first few days of infection.^9^ Recent evidence suggests that whilst the clinical sensitivity of PCR remains very high during the first few days after initial onset of symptoms, it then decreases over time, dropping from >90% over the first 5 days, to 70–71% between days 9–11 and 30% on day 21 following onset of symptoms.^10^ Conversely, the clinical sensitivity of serological tests has been shown to increase over time following initial onset of symptoms, from >50% at day 7, to >80% and then finally 100% at days 12 and 21, respectively, using an in-house enzyme-linked immunosorbent assay (Massachusetts General Hospital, Boston, MA, USA and the Ragon Institute of MGH, MIT and Harvard, Cambridge, MA, USA).^10^ Therefore, it is possible that complementary, time-dependent use of PCR and serological testing will increase reliability when determining prior exposure to SARS-CoV-2 infection, which could better inform morbidity and mortality rates and virus containment measures.

The Elecsys® Anti-SARS-CoV-2 electrochemiluminescence immunoassay (Roche Diagnostics International Ltd, Rotkreuz, Switzerland) was developed to provide an accurate method for the *in vitro* qualitative detection of antibodies to SARS-CoV-2, including IgG, in human serum and plasma, using a recombinant protein representing the N antigen of SARS-CoV-2.^11^ Previously, the Elecsys Anti-SARS-CoV-2 immunoassay demonstrated very high specificity in diagnostic routine (99.9%) and blood donor screening (99.8%) samples collected prior to the onset of the COVID-19 pandemic (pre-September 2019) and thus presumed negative for SARS-CoV-2.^12^ High specificity (99.8%) and sensitivity (99.5%) were also previously observed for the Elecsys Anti-SARS-CoV-2 immunoassay in samples with prior PCR-confirmed SARS-CoV-2 infection, supporting its use as a tool for identification of past SARS-CoV-2 infection.^13^ The aims of this multi-centre study were to further evaluate the sensitivity of the Elecsys Anti-SARS-CoV-2 immunoassay in single and sequential serum and plasma samples with PCR-confirmed past exposure to SARS-CoV-2, to provide broader evidence on assay performance, and compare results versus commercially available comparators.

## Materials and Methods

### Study design

The performance of the Elecsys Anti-SARS-CoV-2 immunoassay was retrospectively evaluated at three diagnostic laboratories in Germany (Augsburg, Heidelberg and Berlin) between 20^th^ May 2020 (first sample tested) and 2^nd^ September 2020 (last sample tested). All study sites provided anonymised, frozen, residual serum or plasma for inclusion in both sensitivity and method comparison analyses. All samples were single or sequential, and confirmed positive for SARS-CoV-2 using PCR. Augsburg and Heidelberg included samples referred to the respective study site by physicians. Heidelberg also included samples from employees of MVZ Labor Limbach and hospitalised patients, including a subset from patients receiving dialysis. All samples provided by the study site in Berlin were collected from hospitalised patients, including a subset from patients monitored in the intensive care unit (ICU). Samples of unknown matrix, with non-removable precipitates or turbidities, a sample volume <300 µL, SARS-CoV-2 PCR confirmation missing or negative, time difference between blood draw and PCR missing or negative, and/or pre-characterised samples with all antibody results confirmed negative were excluded.

The study was conducted in accordance with the study protocol provided by Roche Diagnostics and in accordance with the principles of the Declaration of Helsinki. All human samples utilised at the three study sites in Germany (Augsburg, Heidelberg, Berlin) were anonymised, frozen, residual samples, therefore no ethical approval or waiver was required in accordance with local legislation from ZEKO (Central Ethics Commission at the German Medical Association). A statement was obtained from the Ethics Committee of the Landesärztekammer Bayern confirming that there are no objections to the coherent use of anonymised residual samples.

### Assay

The Elecsys Anti-SARS-CoV-2 electrochemiluminescence immunoassay was developed for the *in vitro* qualitative detection of antibodies to SARS-CoV-2, including IgG, in human serum and plasma. The Elecsys Anti-SARS-CoV-2 immunoassay is intended for use on cobas e analysers, and utilises a recombinant protein representing the N antigen in a double-antigen sandwich test format to detect antibodies to SARS-CoV-2.^14^ The Elecsys Anti-SARS-CoV-2 immunoassay detects antibodies independent of isotype, detecting predominantly mature, high-affinity IgG, but also IgA and IgM antibodies.^11^ The total duration of the immunoassay is 18 minutes, and the analyser automatically calculates a cut-off based on the measurement of two calibrators, one negative (ACOV2 Cal1) and one positive (ACOV2 Cal2). The result of a sample is given as either ‘reactive’ or ‘non-reactive’ in the form of a cut-off index (COI).

In this study, measurements determined using the Elecsys Anti-SARS-CoV-2 immunoassay were interpreted according to the manufacturer’s instructions. Samples with a COI <1.0 were considered non-reactive and deemed negative for anti-SARS-CoV-2 antibodies, while those with a COI ≥1.0 were considered reactive and deemed positive for anti-SARS-CoV-2 antibodies.^14^

### Sensitivity

The sensitivity of the Elecsys Anti-SARS-CoV-2 immunoassay (on cobas e 601 and 801 analysers; Roche Diagnostics International Ltd, Rotkreuz, Switzerland) was evaluated using anonymised, residual, frozen serum or plasma samples from patients with PCR-confirmed SARS-CoV-2 infection. All samples were categorised by the week in which they were drawn following a positive PCR result, and grouped as follows: 0–6 days, 7–13 days, or ≥14 days post-PCR confirmation. For sequential samples, blood draws were performed over a period of minimum 2 to maximum 64 days. If more than one sample per patient was collected per time interval, only the last specimen per patient was included in the sensitivity calculation. For example, if blood was drawn from a patient on day 3, day 7, day 10, day 14 and day 21, only values from day 3, day 10 and day 21 would be included in the 0–6, 7–13 and ≥14 day post-PCR confirmation groups, respectively. Overall and site-specific sensitivity were calculated using samples from patients for whom blood draws were collected ≥14 days post-PCR confirmation of SARS-CoV-2 infection.

### Method comparison

For method comparisons, sensitivity results determined using the Elecsys Anti-SARS-CoV-2 immunoassay (cobas e 601 and 801 analysers) were compared with those calculated for other commercially available SARS-CoV-2 assays, as available at each study site. For a breakdown of comparator assays by study site, please refer to **Supplementary Table 1**. Comparator platforms included the Euroimmun Anti-SARS-CoV-2 IgG assay, Abbott ARCHITECT SARS-CoV-2 IgG assay, Siemens ADVIA Centaur SARS-CoV-2 Total assay, and YHLO iFlash SARS-CoV-2 IgG and IgM assays. Results for each comparator assay were interpreted using cut-off values provided in the respective manufacturer’s package insert.

Sensitivity was calculated for the Elecsys Anti-SARS-CoV-2 immunoassay and each comparator assay, including samples with valid results on both assays only, at time intervals of 0–6, 7–13 and ≥14 days post-PCR confirmation of SARS-CoV-2 infection.

For all comparator assays, results considered ‘equivocal’ or ‘borderline’ per the cut-off values provided in the manufacturer’s package insert were herein grouped into a ‘grey zone’. For assays with results that fell into the ‘grey zone’, two calculations were performed. In the first, all samples with grey zone results were excluded from the analysis. In the second, all grey zone results were interpreted as reactive.

### Sample handling and data management

Transport and storage of all study materials was performed by a third-party vendor (TRIGA-S Scientific Solutions, Habach, Germany). Frozen samples were shipped on dry ice in thermally insulated containers and were received at the destination in a frozen state. Upon receipt at the study site, all samples were stored at –20°C or –80°C until testing. Assay results were obtained from instrument export files. At the end of the study, all samples were disposed of according to the routine clinical requirements of the respective study sites, or shipped to the Roche Diagnostics GmbH (Penzberg, Germany) sample repository for storage.

### Statistical analysis

Sample size estimation for sensitivity analyses was performed using previously published formulae.^15^ It was determined that for an assumed sensitivity of 0.999 a sample size of 32–50 samples would be required to obtain a significance level of 0.05 at a power of 0.8.

For determination of sensitivity, point estimates and two-sided 95% confidence intervals (CIs) were calculated using the exact method in R version 3.4.0.^16^ Acceptance criteria required that 95% CIs for sensitivity overlapped with the CI listed in the Elecsys Anti-SARS-CoV-2 immunoassay method sheet (97.0–100%).^14^ For method comparisons, two-sided Wald CIs were calculated for the differences between estimated sensitivities for the Elecsys Anti-SARS-CoV-2 immunoassay and comparator assays, as recommended by Wenzel and Zapf (2013).^17^ If these CIs did not include zero, differences were considered statistically significant.

## Results

### Analysis set

In total, 806 single and sequential SARS-CoV-2 PCR-confirmed positive samples, collected from 255 patients across the three study sites, were included in this analysis (**Table 1**). Twenty samples were excluded due to an unclear or negative PCR result, missing Elecsys Anti-SARS-CoV-2 immunoassay result, or incorrect inclusion in the sample cohort.

**Table 1.**
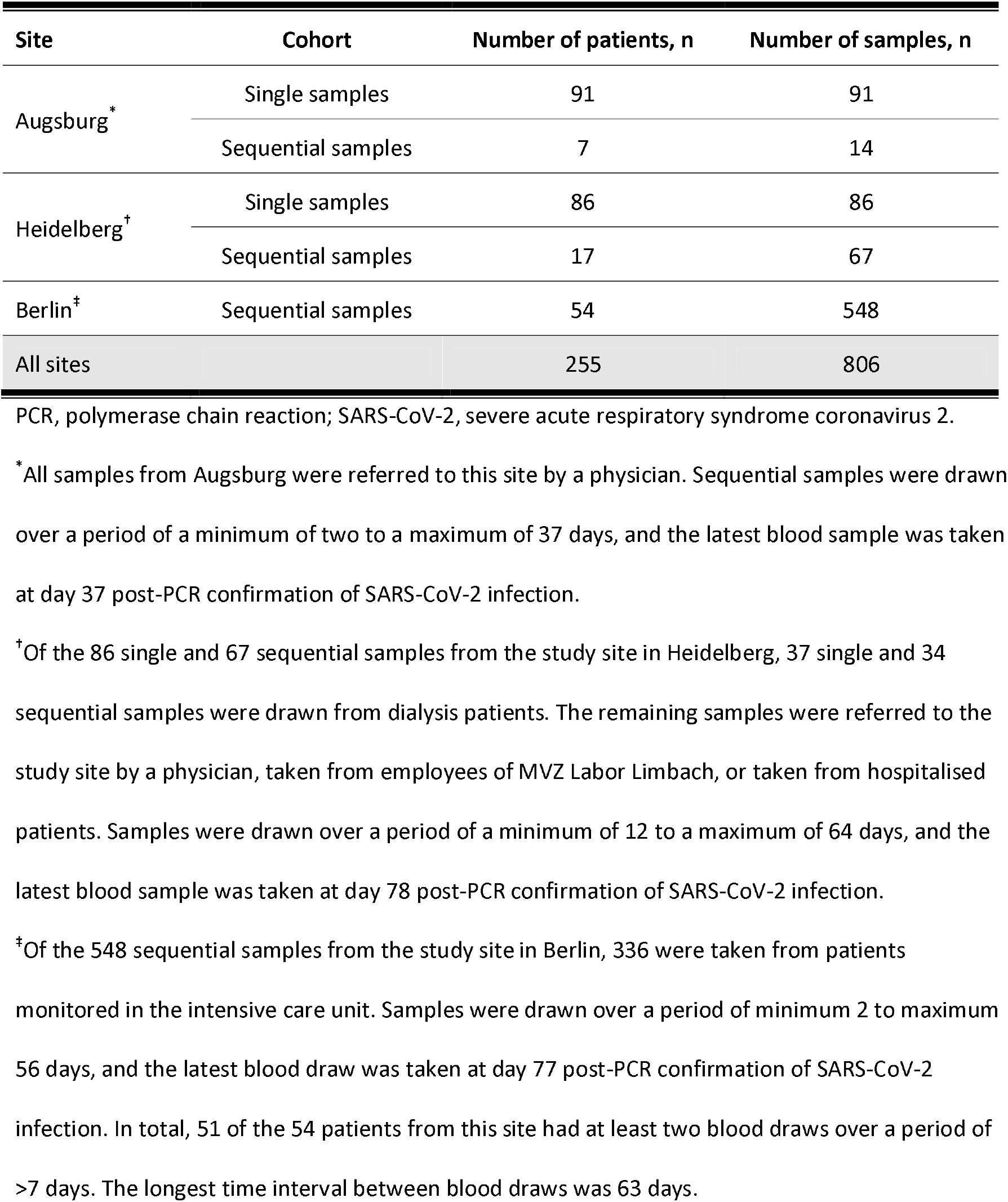
Summary of SARS-CoV-2 PCR-confirmed positive serum and plasma samples by cohort and site

Of the 91 single and 14 sequential samples from seven patients from the study site in Augsburg, all were drawn from patients referred to this site by a physician. Sequential samples were drawn over a period of a minimum of two to a maximum of 37 days, and the latest blood sample was taken at day 37 post-PCR confirmation of SARS-CoV-2 infection.

Of the 86 single and 67 sequential samples from the study site in Heidelberg, 37 single and 34 sequential samples were drawn from dialysis patients. Samples were drawn over a period of a minimum of 12 to a maximum of 64 days, and the latest blood sample was taken at day 78 post-PCR confirmation of SARS-CoV-2 infection.

Of the 548 sequential samples from the study site in Berlin, 336 were taken from patients monitored in the ICU. Samples were drawn over a period of a minimum of two to a maximum of 56 days, and the latest blood sample was taken at day 77 post-PCR confirmation of SARS-CoV-2 infection. In total, 51 of the 54 patients from this site had at least two blood draws over a period of >7 days. The longest time interval between blood draws was 63 days.

### Sensitivity

Overall sensitivity of the Elecsys Anti-SARS-CoV-2 immunoassay was 93.6% (95% CI 89.5–96.5) in samples collected ≥14 days post-PCR confirmation of SARS-CoV-2 infection (n = 219; Table 2). Fourteen samples were found to be non-reactive: 12 from Heidelberg, one from Augsburg and one from Berlin. Across different time intervals for blood draws post-PCR confirmation, sensitivity was highest at 56–62 days (100% [95% CI 85.8–100]) and ≥63 days (100% [95% CI 76.8–100]), and lowest at 0–6 days (43.1% [95% CI 29.3–57.8]).

**Table 2.**
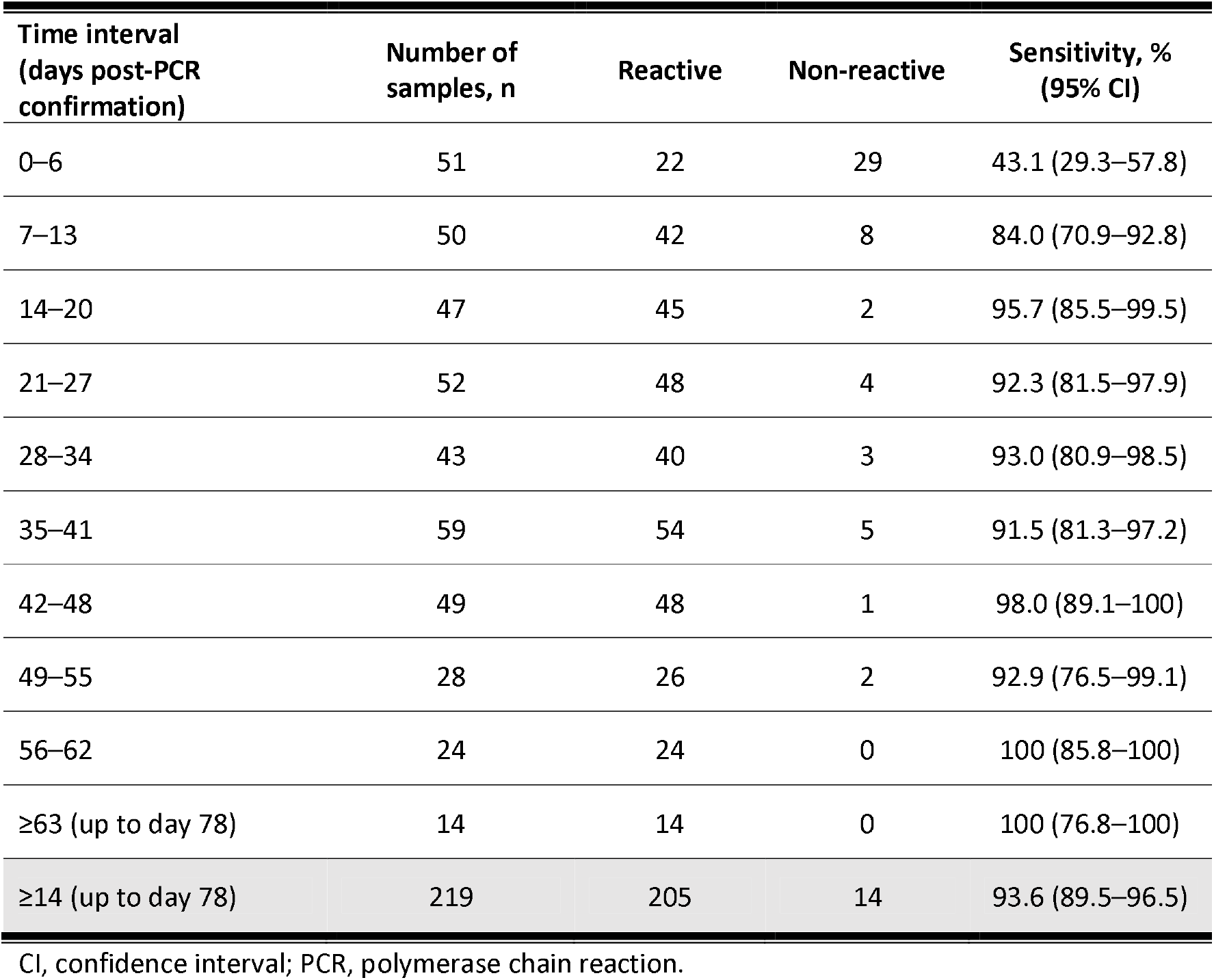
Sensitivity of the Elecsys Anti-SARS-CoV-2 immunoassay by time interval post-PCR confirmation

Site-specific calculations showed that sensitivity of the Elecsys Anti-SARS-CoV-2 immunoassay was highest at Augsburg (98.9% [95% CI 94.2–100]) in samples drawn ≥14 days post-PCR confirmation, and lowest at Augsburg in samples drawn 0–6 days post-PCR confirmation (33.3% [95% CI 0.84–90.6]). For all three sites, sensitivity was highest at ≥14 days post-PCR confirmation and lowest at 0–6 days post-PCR confirmation (**Table 3**).

**Table 3.** Sensitivity of the Elecsys Anti-SARS-CoV-2 immunoassay by time interval and site

| Time interval<br>(days post-PCR<br>confirmation) | Sensitivity by site, % (95% CI) |  |  |  |  |  |
| --- | --- | --- | --- | --- | --- | --- |
|  | Number of<br>samples, n | Augsburg | Number of<br>samples, n | Heidelberg | Number of<br>samples, n | Berlin |
| 0–6 | 3 | 33.3<br>(0.84–90.6) | 25 | 40.0<br>(21.1–61.3) | 23 | 47.8<br>(26.8–69.4) |
| 7–13 | 5 | 80.0<br>(28.4–99.5) | 11 | 81.8<br>(48.2–97.7) | 34 | 85.3<br>(68.9–95.0) |
| ≥14<br>(up to day 78) | 94 | 98.9<br>(94.2–100) | 84 | 85.7<br>(76.4–92.4) | 41 | 97.6<br>(87.1–99.9) |

CI, confidence interval; PCR, polymerase chain reaction.

### Method comparison

Method comparison was performed using 11–124 samples per comparison across the three study sites for the time intervals 0–6 days, 7–14 days and ≥14 days post-PCR confirmation. Across all assays evaluated, sensitivity was highest for the Euroimmun Anti-SARS-CoV-2 IgG assay (with grey zone values included and considered reactive) for samples drawn ≥14 days post-PCR confirmation at 95.2% (95% CI 89.8–98.2; **Table 4**).

**Table 4.**
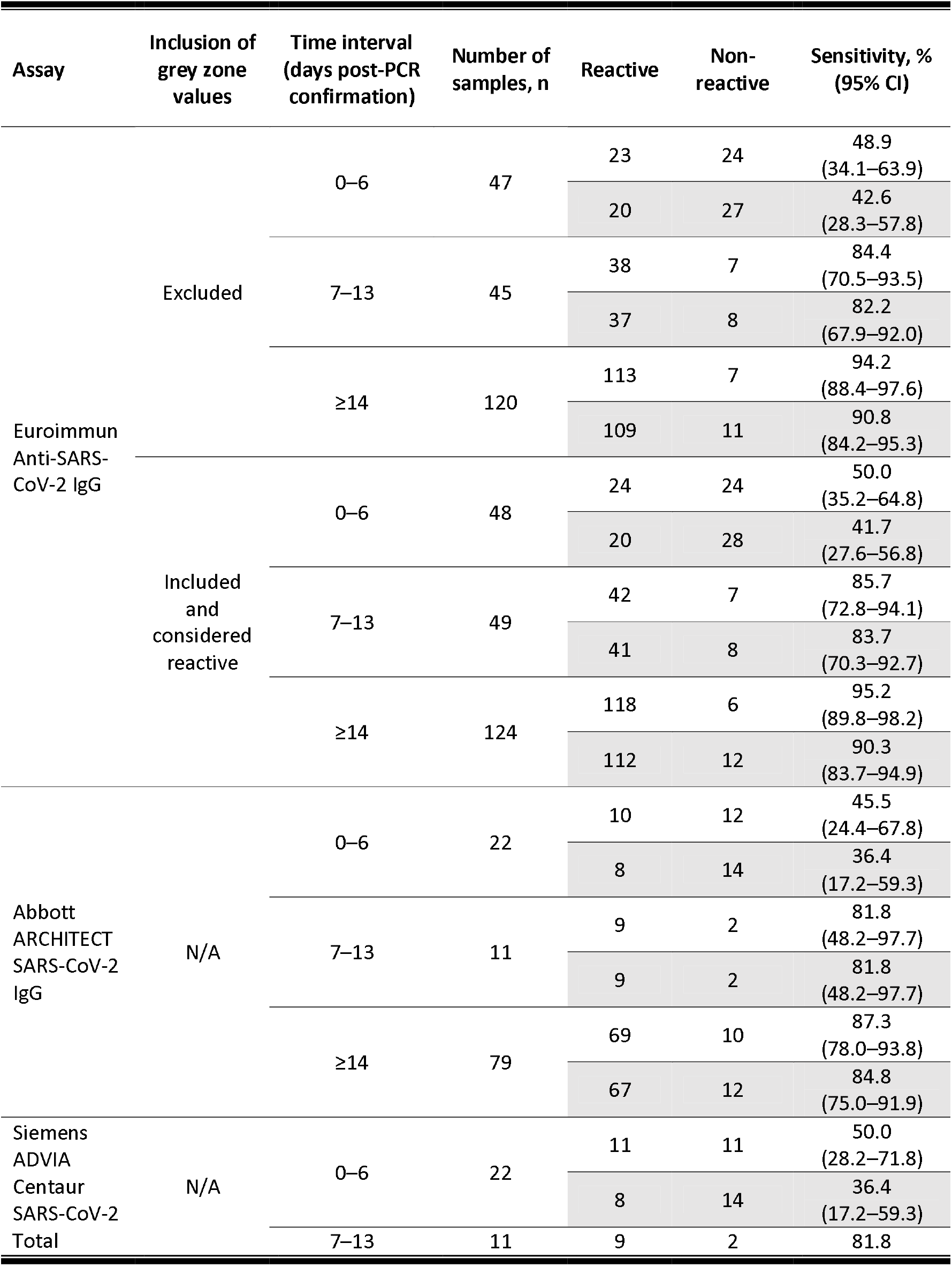
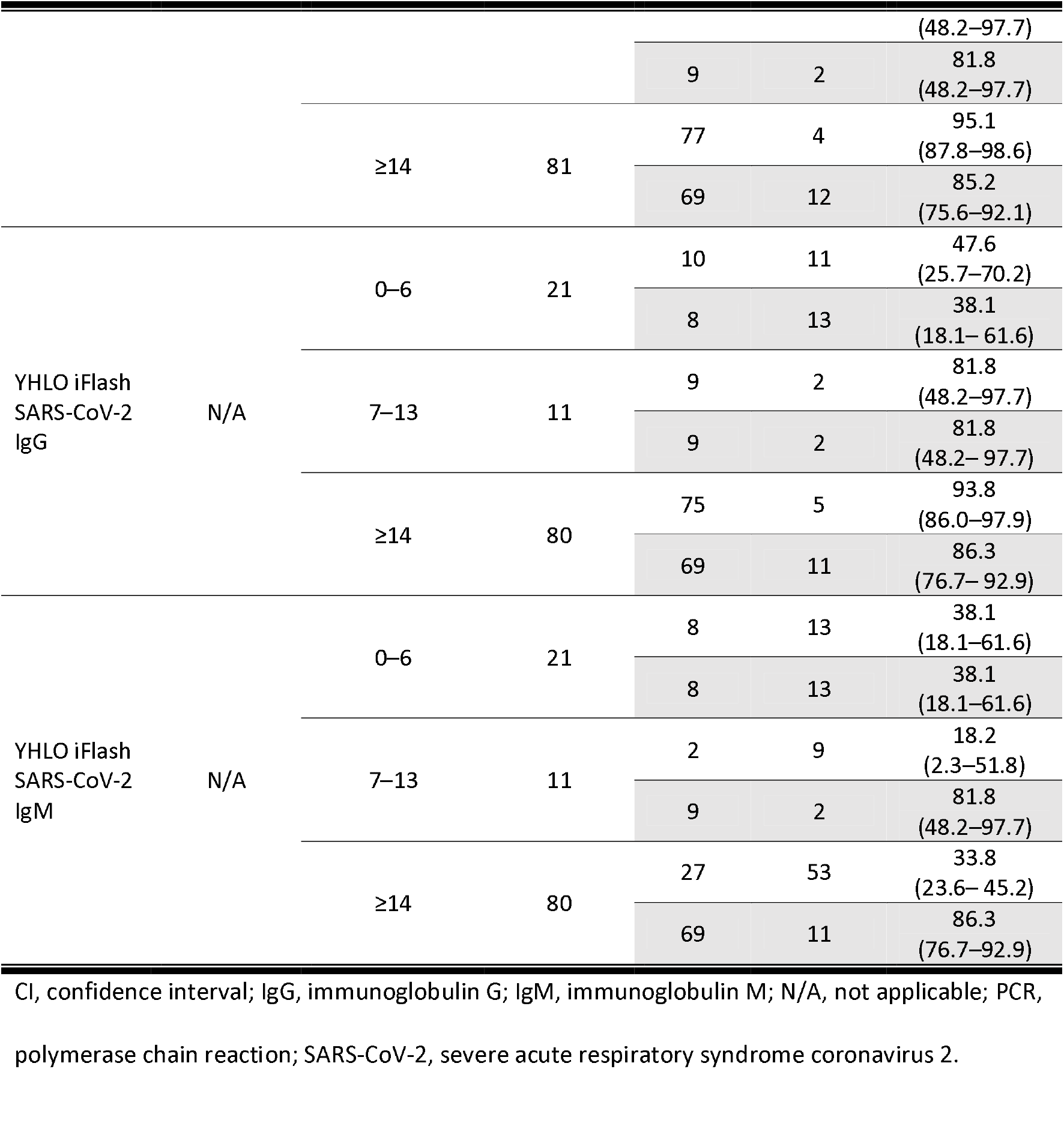
Summary of sensitivity values for the Elecsys Anti-SARS-CoV-2 immunoassay and comparator assays by time interval across all sites Measurements taken on the Elecsys Anti-SARS-CoV-2 immunoassay using the same samples are shaded in grey.

Sensitivity was comparable between the Elecsys Anti-SARS-CoV-2 immunoassay and the Euroimmun Anti-SARS-CoV-2 IgG (n = 124, 90.3% [95% CI 83.7–94.9] versus 95.2% [95% CI 89.8– 98.2], difference: −4.84% [95% CI −8.62–1.39]) and Abbott ARCHITECT SARS-CoV-2 IgG (n = 79, 84.8% [95% CI 75.0–91.9] versus 87.3% [95% CI 78.0–93.8], difference: −2.53% [95% CI −9.53–4.46]) assays for samples drawn ≥14 days post-PCR confirmation. No statistically significant differences were observed between these assays.

Both the Siemens ADVIA Centaur SARS-CoV-2 Total and the YHLO iFlash SARS-CoV-2 IgG assays were found to have significantly higher sensitivity compared with the Elecsys Anti-SARS-CoV-2immunoassay for samples drawn ≥14 days post-PCR confirmation (n = 81, 95.1% [95% CI 87.8–98.6] versus 85.2% [95% CI 75.6–92.1], difference: 9.88% [95% CI 1.08–18.7]; n = 80, 93.8% [95% CI 86.0– 97.9]; versus 86.3% [95% CI 76.7–92.9], difference: 7.50% [95% CI 0.768–14.2]). In addition, the Elecsys Anti-SARS-CoV-2 immunoassay demonstrated significantly higher sensitivity compared with the YHLO iFlash SARS-CoV-2 IgM assay for samples drawn ≥14 days post-PCR confirmation (n = 80, 86.3% [95% CI 76.7–92.9] versus 33.8% [95% CI 23.6–45.2], difference: 52.5% [95% CI 38.9–63.1]).

## Discussion

The COVID-19 pandemic created an urgent unmet need for highly sensitive serological assays capable of detecting antibodies to SARS-CoV-2, in order to aid identification of individuals previously exposed to the virus and inform containment procedures.^18,19^ The present findings indicate that the Elecsys Anti-SARS-CoV-2 immunoassay demonstrates high overall sensitivity in samples collected ≥14 days post-PCR confirmation of SARS-CoV-2 infection (93.6% [95% CI 89.5–96.5]) across multiple sites, providing broader evidence for the favourable performance of the assay across different diagnostic laboratories, and supporting its use in routine screening for previous exposure to SARS-CoV-2.

The 95% CI reported here for overall sensitivity of the Elecsys Anti-SARS-CoV-2 immunoassay (93.6% [95% CI 89.5–96.5]) did not overlap with the 95% CI reported in the manufacturer’s package insert (99.5% [95% CI 97.0–100]^14^) when data from all three sites were combined. Taken separately, 95% CIs for sensitivity of the Elecsys Anti-SARS-CoV-2 immunoassay from both Augsburg (98.9% [95% CI 94.2–100]) and Berlin (97.6% [95% CI 87.1–99.9]) did overlap with those provided in the manufacturer’s package insert for samples collected ≥14 days post-PCR confirmation, but 95% CIs calculated for Heidelberg (85.7% [95% CI 76.4–92.4]) did not. However, it is important to consider that the majority of samples utilised at the study site in Berlin were taken from patients in the ICU, for whom blood draws were closely monitored. In contrast, the majority of samples tested at Augsburg and Heidelberg were from patients referred to each respective site by a physician. For patients in the ICU, disease severity was likely far greater than that of non-hospitalised patients infected with SARS-CoV-2, which may exacerbate immune response and increase the likelihood of a positive test result from the Elecsys Anti-SARS-CoV-2 immunoassay. This is supported by findings from Guthmiller et al. (2020), who observed that patients with an increased humoral response against the viral S and N proteins exhibit more severe symptoms of SARS-CoV-2 infection.^20^ Other studies have also reported that the early IgG response in severely ill patients is stronger compared with non-severe cases; however, there remains some discussion regarding these results.^21,22^ It is possible that the observed difference in sensitivity performance of the Elecsys Anti-SARS-CoV-2 immunoassay among sites was caused in part by the pre-characterisation and selection of cohorts applied at each study site; however, given the unprecedented situation of the ongoing COVID-19 pandemic, the clinical heterogeneity included in this study should be considered a strength rather than a limitation.

Moreover, approximately 50% of all samples tested at Heidelberg were from patients undergoing dialysis, in whom infections are one of the main causes of morbidity and mortality.^23,24^ Patients with end-stage renal disease (ESRD) have poor renal function, which is associated with uraemia-associated immune deficiency.^24^ In addition, it has been shown that even a single dialysis procedure can have an immunomodulatory effect, contributing to immune deficiency in patients with ESRD.^25^ Therefore, it is less likely that these patients were able to mount a detectable antibody response to SARS-CoV-2.^26,27^ This could explain the fact that 10 of 12 non-reactive samples collected ≥14 days post-PCR confirmation at Heidelberg were from patients receiving dialysis, and supports the use of PCR confirmation alongside serological testing for immunocompromised patients in order to ensure an accurate test result.

In accordance with previous findings, the sensitivity of the Elecsys Anti-SARS-CoV-2 immunoassay was lowest 0–6 days post-PCR confirmation at 43.1%, before increasing to >90% for samples drawn ≥14 days post-PCR confirmation.^10,13^ This trend towards increased sensitivity over time following PCR confirmation was observed across all three sites, with sensitivity ranging from 33.3–47.8% for samples drawn 0–6 days post-PCR confirmation, 80.0–85.3% for samples drawn 7–13 days post-PCR confirmation and 85.7–98.9% for samples drawn ≥14 days post-PCR confirmation. It was also observed across all comparator assays except for the YHLO iFlash SARS-CoV-2 IgM assay, for which sensitivity was highest for samples drawn 0–6 days post-PCR confirmation (38.1%), and lowest in samples drawn 7–13 (18.2%) and ≥14 (33.8%) days post-PCR confirmation. However, in a similar cohort of SARS-CoV-2 PCR-confirmed positive samples, Oved et al. (2020) previously found that approximately 5% of patients remained seronegative ≥14 days post-PCR confirmation, and thus did not seroconvert.^28^ It is important to consider this phenomenon when evaluating these results, as it could affect the trends observed herein.

The low sensitivity of the YHLO iFlash SARS-CoV-2 IgM assay is in accordance with that observed previously by Kittel et al. (2020).^29^ A potential explanation for this is the fact that detection of SARS-CoV-2-specific IgM is limited to very early in the infection cascade; only 20% of SARS-CoV-2 infected individuals present IgM before IgG, and the majority will present both IgM and IgG in tandem.^30^ Some studies have reported the detection of SARS-CoV-2-specific IgG even before IgM.^31,32^ Taken together, the clinical value of IgM for diagnosis of COVID-19 remains unclear.^33^

Method comparisons demonstrated that the Elecsys Anti-SARS-CoV-2 immunoassay had significantly higher sensitivity than the YHLO iFlash SARS-CoV-2 IgM assay (86.3% versus 33.8%) in this cohort, whilst also demonstrating comparable sensitivity to several other commercially available assays, including the Euroimmun Anti-SARS-CoV-2 IgG (90.3% versus 95.2%) and the Abbott ARCHITECT SARS-CoV-2 IgG (84.8% versus 87.3%) assays. However, it is important to consider the different antibodies targeted by each assay when drawing direct comparisons between them. For example, the format of the Elecsys Anti-SARS-CoV-2 immunoassay requires binding of an antibody in the patient sample to two specific antigens, and as such favours preferential detection of mature, high-affinity antibodies characteristic of the late stages of SARS-CoV-2 infection.^11,14^ In addition, the clinical performance of serological assays is a compromise between sensitivity and specificity; whilst some assays are designed for higher sensitivity, others are designed for higher specificity. In future, a comprehensive comparison of both parameters is required.

The sensitivity of the YHLO iFlash SARS-CoV-2 IgM and Abbott ARCHITECT SARS-CoV-2 IgG assays were comparable to previously reported values in SARS-CoV-2 confirmed positive samples collected 13 days and 28–56 days post-symptom onset (42% and 64.5–100%, respectively).^34-37^ Previously, higher sensitivity for the YHLO iFlash SARS-CoV-2 IgG assay was reported in COVID-19 confirmed positive samples compared with present findings (76.9–94.0%).^35,37^ Interestingly, sensitivity for the Euroimmun Anti-SARS-CoV-2 IgG assay reported here was higher compared with that of previous studies in COVID-19 confirmed samples (65.0–78.0%).^37,38^

A major strength of this study was the large cohort of PCR-confirmed SARS-CoV-2 positive samples tested in a routine setting across multiple sites and patient groups, ensuring that reliability of the point estimates remained high. This study was performed under accelerated timelines due to the high scientific value of the data in the ongoing COVID-19 pandemic, and so long-term stability data for frozen samples was not available prior to study initiation; however, the prolonged stability of IgG antibodies is well-documented, and so this should not impact the present data.^39,40^

## Conclusion

The Elecsys Anti-SARS-CoV-2 immunoassay demonstrated a high overall sensitivity of 93.6% (95% CI 89.5–96.5) in samples collected ≥14 days post-PCR confirmation of SARS-CoV-2 infection. Calculated sensitivity was also comparable to several commercially available comparator assays across multiple sites, supporting the use of this immunoassay as a tool to aid in determination of previous exposure to SARS-CoV-2.

## Supporting information

STARD 2015 Checklist

## Data Availability

Qualified researchers may request access to individual patient level data through the clinical study
data request platform (https://vivli.org/). Further details on Roche's criteria for eligible studies are available here: https://vivli.org/members/ourmembers/. For further details on Roche's Global Policy
on the Sharing of Clinical Information and how to request access to related clinical study documents,
see here: https://www.roche.com/research_and_development/who_we_are_how_we_work/clinical_trials/our_commitment_to_data_sharing.htm

## Acknowledgements

The authors would like to acknowledge Kathrin Schoenfeld (Roche Diagnostics) for her role in study conceptualisation, study management, interpretation of analysis and further critical input; Michael Laimighofer (Roche Diagnostics) for his role in database generation and data validation, statistical analysis plan and formal analysis; and Sigrid Reichhuber, Janina Edion, and Yvonne Knack (Roche Diagnostics) for their role in investigational site management, data acquisition and study monitoring. Third-party medical writing support, under the direction of the authors, was provided by Chloe Fletcher, MSc (Ashfield MedComms, an Ashfield Health company, Macclesfield, UK) and was funded by Roche Diagnostics International Ltd, Rotkreuz, Switzerland. COBAS, COBAS E and ELECSYS are trademarks of Roche. All other product names and trademarks are the property of their respective owners.

