## Supplementary materials for "Sensitivity of the Elecsys Anti-SARS-CoV-2 immunoassay as an aid in determining previous exposure to SARS-CoV-2"

**Supplementary Table 1.** Anti-SARS-CoV-2 assays utilised for the method comparison analysis by site

| **Assay^*^** | **Antigen** | **Site** | | |
| --- | --- | --- | --- | --- |
|  |  | **Augsburg** | **Heidelberg** | **Berlin** |
| Euroimmun Anti-SARS-CoV-2 IgG | Spike protein | X | X | X |
| Abbott ARCHITECT SARS-CoV-2 IgG | Nucleocapsid |  | X |  |
| Siemens ADVIA Centaur SARS-CoV-2 Total | Spike protein |  | X |  |
| YHLO iFlash SARS-CoV-2 IgG | Spike protein/ nucleocapsid |  | X |  |
| YHLO iFlash SARS-CoV-2 IgM | Spike protein/ nucleocapsid |  | X |  |

*Eligible samples had test results from both the Elecsys Anti-SARS-CoV-2 immunoassay and the comparator assay. IgG, immunoglobulin G; IgM, immunoglobulin M; SARS-CoV-2, severe acute respiratory syndrome coronavirus 2.
